# Japan’s Covid mitigation strategy and its epidemic prediction

**DOI:** 10.1101/2021.05.06.21256476

**Authors:** Yasuharu Tokuda, Toshikazu Kuniya

## Abstract

The COVID-19 epidemic curve in Japan was constructed based on daily reported data from January 14, 2020 until April 20, 2021. A SEIR compartmental model was used for the curve fitting by updating the estimation per wave. In the current vaccination pace of 1/1000, restrictions (state of emergency in Japan) would be repeated 4 times until the end of next March. In the case of 1/500, another round of restriction would be required in the summer 2021, after which the infection would be mitigated. In the case of 1/250, there would be no need for restriction after the current spring restriction. The scenario of completing the vaccination of 110 million people by the end of March 2020 corresponds to the case of 1/250 in this curve. When considering the likely spread of variant with greater infectiousness (here we assume 1.3 times greater than the original virus), 1/500 pace of vaccination would not be enough to contain it and need several series of restrictions. There are currently several variants of concern that are already spreading in urban areas in this country. In the new stage of the replacement of variants, if the vaccination pace could not be quadrupled from the current pace, Japan could not become a zero covid (zero corona) country at least one year.

## TEXT

We agree with Oliu-Barton M, et al (1) that countries opted for Zero Covid have the lower number of COVID-19 death rate with less economic damage. Japan was included into one of the Organisation for Economic Co-operation and Development (OECD) countries that aim for elimination. However, unfortunately, the government COVID-19 committee of Japan keeps mitigation strategy, so-called With-Corona policy (2). In fact, as of April 26, the cumulative deaths per 100,000 people was 7.9 in Japan and it was 3.5, 26, and 158 times greater than those of South Korea, China, and Taiwan, respectively (3). Regarding annual GDP in 2020 compared that in 2019, Japan is projected to have contracted 5.3%, while South Korea shrank only 1% and China expanded 2.3% (4).

Moreover, we constructed the COVID-19 epidemic curve (Figure 1A) in Japan based on daily reported data from January 14, 2020 until April 20, 2021. The following SEIR compartmental model as in (5,6) was used for the curve fitting.

**Figure 1:**
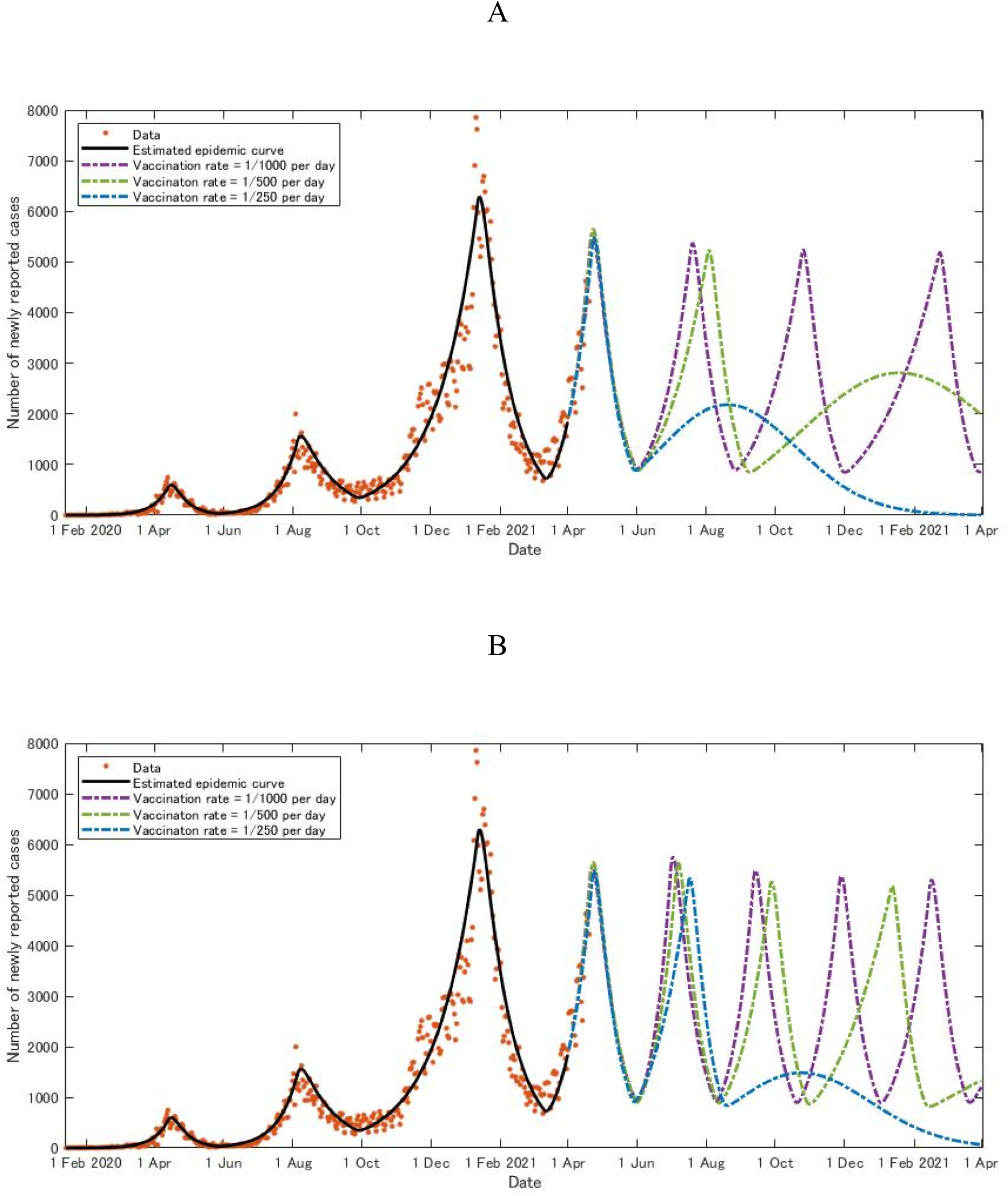
Prediction of COVID-19 by epidemic curve in Japan. A: based on previous data on the infectiousness B: on the increased (1.3 times) infectiousness over the previous data.

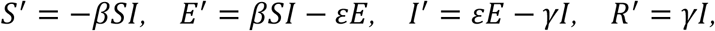

where *S* = *S* (*t*), *E* = *E* (*t*), *I* = *I* (*t*) and *R* = *R* (*t*) represent the susceptible, exposed, infected and removed/recovered populations at time *t*, respectively. The time unit is a day. *β*, *ε* and *γ* represent the infection rate, the transition rates from *E* to *I* and *I* to *R*, respectively. As in (5,6), we assumed that *ε* = 0.2 and *γ* 0.1 so that the average incubation and infection period are 1/*ε* = 5 (days) and 1/*γ* = 10 (days), respectively (7,8). In addition, we set *S* + *E* + *I* + *R* = 1 so that each population is the proportion to the total population. As in (5,6), to estimate the infection rate *β*, we assumed that the daily number of newly reported cases in Japan is given by *Y* = *Y*(*t*) *pI*(*t*)*N* and fitted it to the data (9) to find the best *β* that minimizes the least-squares error, where *p* denotes the detection rate and *N* denotes the population in Japan. We assumed that *N* = 1.26 × 10^8^ (10) and estimated *p* 0.33 by using a similar procedure as in (6). In this study, to estimate the infection rate *β*, we divided the period January 14, 2020 – April 20, 2021 into the following seven parts (see also the actual data in Figure 1A):

(P1) January 14 - April 13, 2020 (the first period of infection spread)

(P2) April 14 - May 25, 2020 (the first period of infection control)

(P3) May 26 - August 4, 2020 (the second period of infection spread)

(P4) August 5 - September 25, 2020 (the second period of infection control)

(P5) September 26, 2020 - January 10, 2021 (the third period of infection spread)

(P6) January 11 - March 10, 2021 (the third period of infection control)

(P7) March 11 - April 20, 2021 (the fourth period of infection spread)

The infection rate *β* was estimated for each period by fitting the model solution to the data. More precisely, we obtained the following estimated values for *β* in each period:

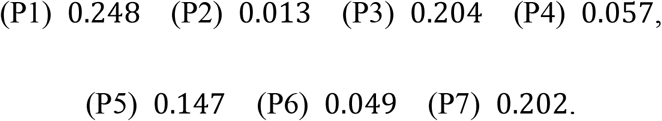

Define *β*^+^ 0.24 and *β*^-^ 0.04 by the average of the above estimated *β* in (P3), (P5), (P7) (the period of infection spread) and (P2), (P4), (P6) (the period of infection control), respectively. Assumptions for prediction after April 21, 2020 include the following: the infection rates in the period of infection spread and control are given by *β*^+^ and *β* ^-^, respectively. If the number of new infections exceeds 5,000 during the period of infection spread, restrictions (lockdown in Japan) will begin. Restrictions will be lifted when the number of new infections falls below 1,000 during the period of infection control. To discuss the effect of the vaccination, we introduced the vaccination rate into the model as follows:

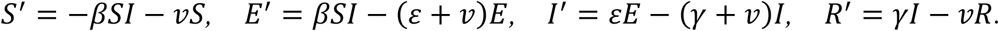

We used three scenarios of vaccination rate: 1/1000 (the current pace), 1/500, 1/250 of unvaccinated persons per day.

As a result, in the pace of 1/1000, the restrictions would be repeated 4 times until the end of next March. In the case of 1/500, another round of restriction would be required in the summer 2021, after which the infection would be mitigated. In the case of 1/250, there would be no need for restriction after the current spring restriction. The scenario of completing the vaccination of 110 million people by the end of March 2020 corresponds to the case of 1/250 in this curve.

However, when considering the likely spread of variant with greater infectiousness (here we assume 1.3 times greater than the original virus), 1/500 pace of vaccination would not be enough to contain it and need several series of restrictions (Figure 1B). As of May 2021, there are several variants of concern including B.1.1.7. that are already spreading in urban areas in this country. Thus, Japan is not currently a member in the Zero Covid community. In this new stage of the replacement of variants, if the vaccination pace could not be quadrupled from the current pace, Japan could not become a member in the community, in which many countries of East Asia and Western Pacific regions belong to, at least one year.

## Data Availability

It is available upon request.

